# Transmission Dynamics of and Insights from the 2018-2019 Measles Outbreak in New York City: A Modeling Study

**DOI:** 10.1101/19005298

**Authors:** Wan Yang

## Abstract

In 2018-2019, New York City experienced the largest measles outbreak in the US in nearly three decades. To identify key factors contributing to this outbreak to aid future public health interventions, here we developed a model-inference system to infer the transmission dynamics of measles in the affected community, based on incidence data. Our results indicate that delayed vaccination of young children aged 1-4 years enabled the initial spread of measles and that increased infectious contact among this age group, likely via gatherings intended to expose unvaccinated children (i.e. “measles parties”), further aggravated the outbreak and led to widespread of measles beyond this age group. We found that around half of infants were susceptible to measles by age 1 (the age-limit to receive the first vaccine dose in the US); as such, infants experienced a large number of infections during the outbreak. We showed that without the implemented vaccination campaigns, the outbreak severity including numbers of infections and hospitalizations would be 10 times higher and predominantly affect infants and children under 4. These results suggest that recommending the first vaccine dose before age 1 and the second dose before age 4 could allow pro-vaccine parents to vaccinate and protect infants and young children more effectively, should high level of vaccine hesitancy persist. In addition, enhanced public health education is needed to reduce activities that unnecessarily expose children to measles and other infections.

## Introduction

Measles is highly contagious and severe viral disease. Thanks to a highly effective vaccine and high coverage of vaccination, endemic transmission of measles—i.e. continuous transmission for more than 12 months—in the US was declared eliminated in 2000. However, due to vaccine hesitancy and declining vaccination rate, in recent years there have been an increasing number of large outbreaks following introduction of measles infection (*1*). Due to long-term fluctuations in vaccine coverage and infection history, population susceptibility could vary substantially by age group. This susceptibility disparity by age can further interact with age-specific social connectivity (i.e., contact rate) to shape the epidemic trajectory. As such, understanding these detailed population characteristics as well as their impact on transmission dynamics in the recent outbreaks is important for devising timely and effective intervention strategies.

In the fall of 2018, several New York City (NYC) residents acquired measles while traveling abroad and subsequently led to the largest measles outbreak in the US in nearly three decades. The first case of this outbreak developed a rash on Sep 30, 2018 and as of Aug 6, 2019 (at the time of this writing), there have been 642 confirmed cases, largely occurring in an Orthodox Jewish community (*2, 3*). To contain the outbreak, the NYC Department of Health and Mental Hygiene (DOHMH) launched extensive vaccination campaigns and, on Apr 9, 2019, ordered mandatary vaccination of all individuals living, working or going to school in the affected zip codes. As a result, over 32 thousand individuals under 19 years were vaccinated with the measles, mumps, and rubella (MMR) vaccine during Oct 2018 – July 2019 and the outbreak subsided (*3, 4*).

In this study, we model the transmission dynamics of this measles outbreak in the affected Orthodox Jewish community in NYC from Oct 1, 2018 to July 31, 2019, months with more than one measles cases reported. Using an age-structured model-inference framework, we are able to estimate key epidemiological features including the initial susceptibilities in five different age groups (i.e., <1, 1-4, 5-17, 18-49, and 50+ years) and the basic reproductive number *R*_*0*_, infer key factors contributing to the spread of measles, estimate the proportions of infection attributable to each age group, as well as assess the impact of vaccination campaigns. We also discuss the implications of our findings to current measles vaccination policies.

## Results

### Overview of the measles outbreak and model fit

The measles outbreak started on Sep 30, 2018, when a young child developed a rash. It evolved relatively slowly in the first three months; however, the outbreak took off quickly in early 2019, peaked in April after the city declaring a public health emergence, and recorded a total of 642 cases by July 31, 2019. As shown in Fig. 1, age-grouped incidence, estimated based on health reports/alerts (*3, 5*), peaked first in Mar 2019 among 1-4 year-olds—the age group with the largest number of infection (275 cases or 42.8% of the total, as of Aug 6, 2018), followed by <1 year-olds (100 cases or 15.6%) and 5-17 year-olds (138 cases or 21.5%) in Apr 2019, and 18+ year-olds (129 cases or 20.1%) in May 2019.

**Fig. 1.**
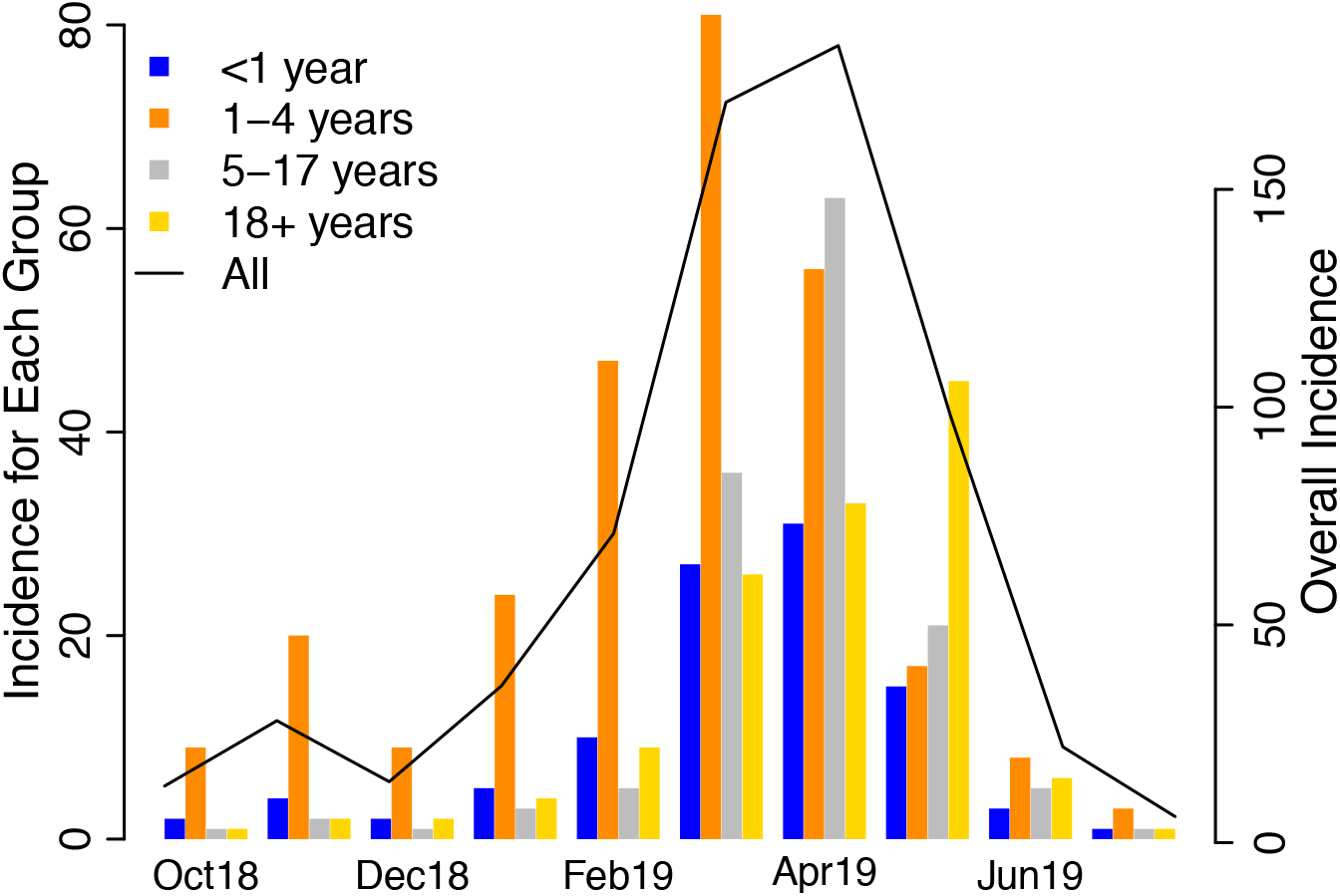
Monthly incidence for all ages and by age group. The solid line (y-axis on the right) shows monthly incidence for all ages, reported as of Aug 6, 2019. For comparison, bars (y-axis on the left) show monthly incidence for <1 (blue), 1-4 (orange), 5-17 (grey) and 18+ (yellow) year-olds, respectively, estimated based on health reports.

As shown in Fig. 2 and Table S2, our model-filter system was able to recreate the overall incidence curve during Oct 2018 – July 2019, estimate the overall age distribution of measles cases, as well as recreate the estimated age-grouped incidence curves for all age groups. Note that while our model (Eqn 1) divided 18+ year-olds into two subgroups (i.e. 18-49 and 50+ years) given their differences in contact rates and interactions with other age-groups, we present the combined results here because data were only available for the entire age group. The estimated reporting rate was around 90% throughout the study period and slightly lower in Apr 2019, at the peak of the outbreak [89.1%, 95% credible interval (CI): 79.4, 99.2%; Fig. S2].

**Fig. 2.**
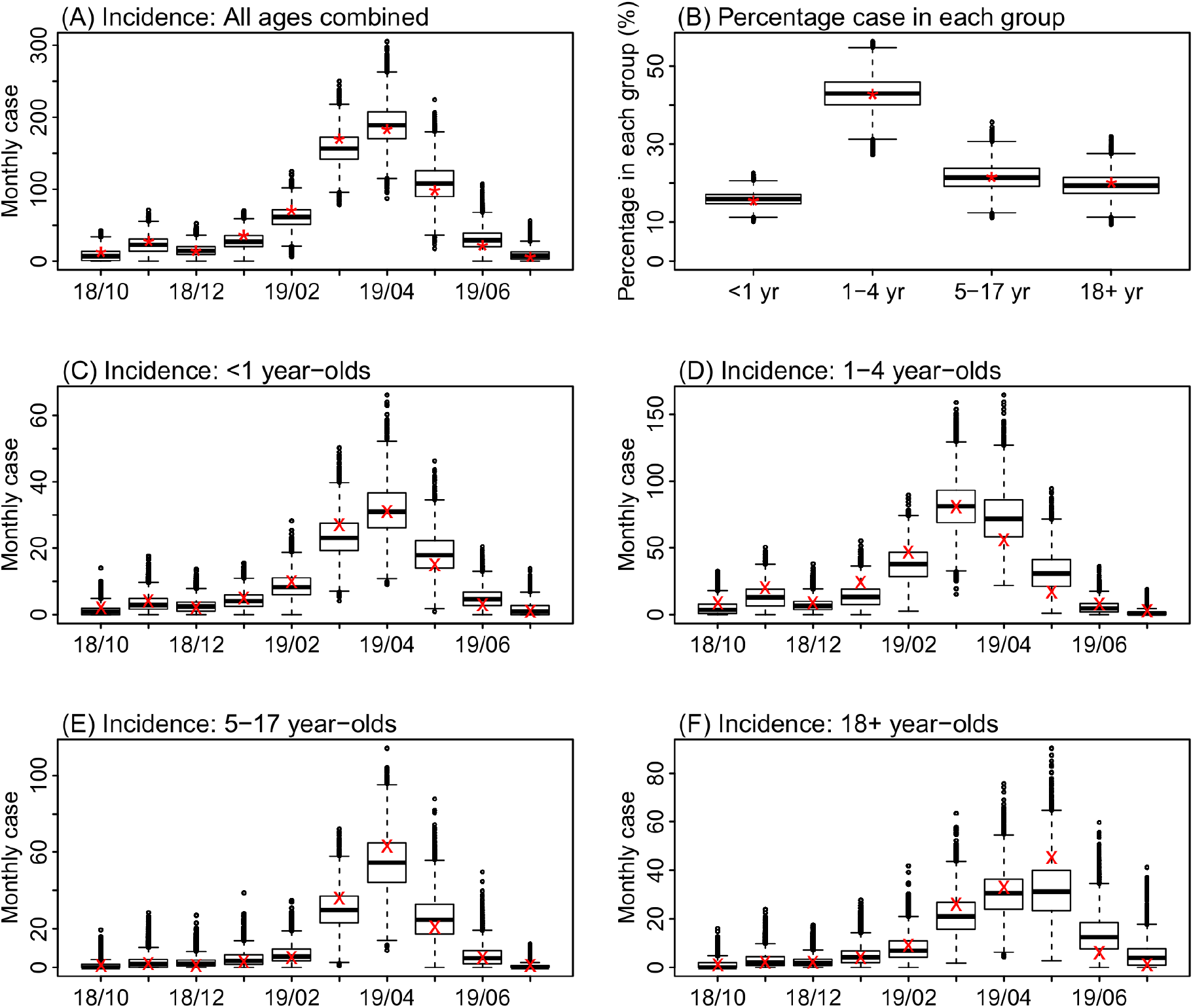
Model fit. Box plots show estimates of monthly incidence for all ages (A), percentage of cases reported in each age group (B), and monthly incidence for <1 year-olds (C), 1-4 year-olds (D), 5-17 year-olds (E), and 18+ year-olds (F). Results are pooled over all 10 model-filter runs (each with 10,000 and in total 100,000 model realizations). Horizontal thick lines show the median of model estimates; box edges show the 25^th^ and 75^th^ percentiles; whiskers show the 2.5^th^ and 97.5^th^ percentiles; and dots show outliers. Stars (*) in A and B show monthly incidence for all ages and the age distribution, reported as of Aug 6, 2019; crosses (x) in C-F show age-grouped monthly incidence estimated from health reports.

### Inference of key epidemiological characteristics

The model-filter system estimated that, at the beginning of the outbreak (i.e. Sep 2018), susceptibility was the highest in infants, at approximately 53.2% (95% CI: 49.0, 57.5%; Fig. 3A). This was expected, because maternal immunity wanes within 3 to 9 months after birth and, as a result, by their first birthday—the age eligible to receive the first dose of MMR vaccine in the US—almost all infants have lost their maternal immunity and are susceptible to measles. Young children aged 1-4 years had the second highest susceptibility; approximately 24.9% (95% CI: 20.4, 29.7%) were susceptible. In comparison, susceptibility was lower among both 5-17 year-olds (6.0%, 95% CI: 4.1, 7.9%) and 18+ year-olds 6.0% (95% CI: 4.4, 7.6%; Fig. 3). These estimates were consistent with the observation that all cases recorded in Oct 2018 were children ranging from 11 months to 4 years (*2*). Sensitivity analysis on assumptions related to the vaccination campaigns showed that estimated susceptibilities were slightly higher for 5-18 year-olds (7.5%, 95% CI: 5.1, 9.9%) and 18-49 year-olds (7.9%, 95% CI: 5.2, 9.9%) if more vaccine doses were given to the Jewish Orthodox community or 5-18 year-olds; however, the estimates were in general consistent with the baseline scenario (Table S2).

**Fig. 3.**
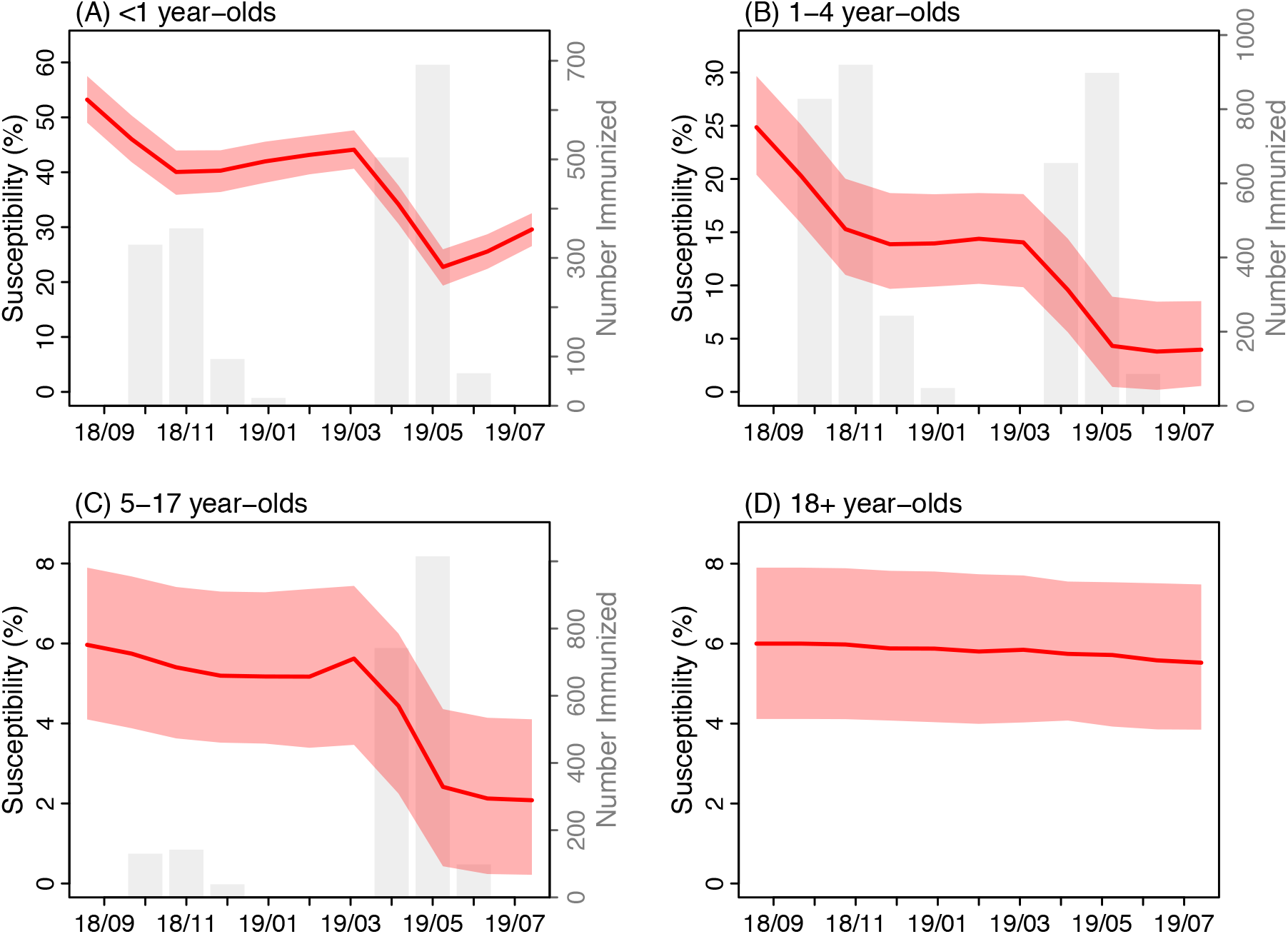
Estimated changes in population susceptibility. Red lines and surrounding areas (y-axis on the left) show the mean and 95% credible intervals of estimates pooled over all 10 model-filter runs (100,000 model realizations in total) for <1 year-olds (A), 1-4 year-olds (B), 5-17 year-olds (C) and 18+ year-olds (D), respectively, at the end of each month from Sep 2018 to July 2019. The initial susceptibilities, estimated at the end of Sep 2018, were computed by adding the total numbers of individuals immunized by the vaccination campaigns in Oct 2018 to the posterior estimates at the end of Oct 2018. For comparison, the grey bars (y-axis on the right) show estimated numbers of individuals immunized during the vaccination campaigns; note that the vaccination campaigns targeted individuals under 19 years and thus is not shown for 18+ year-olds.

The initial vaccination campaigns launched promptly afterwards lowered the susceptibility to 40.3% (95% CI: 36.4, 44.0%) in infants and 13.9% (95%: 9.7, 18.7%) in 1-4 year-olds, by the end of Dec 2018 (Fig. 3). These efforts along with other transmission and infection controls (*2*) appeared to effectively contain the outbreak at the time. The effective reproductive number (*R*_*e*_) is a key epidemiological parameter reflecting the potential of an infection to cause an epidemic in a partially immune population; an epidemic is possible when *R*_*e*_>1. The estimated *R*_*e*_ was 1.5 (95% CI: 0.7, 2.9) in Oct 2018 and dropped to round 1 (95% CI: 0.7, 1.5) in Dec 2018 (Fig. 4B).

**Fig. 4.**
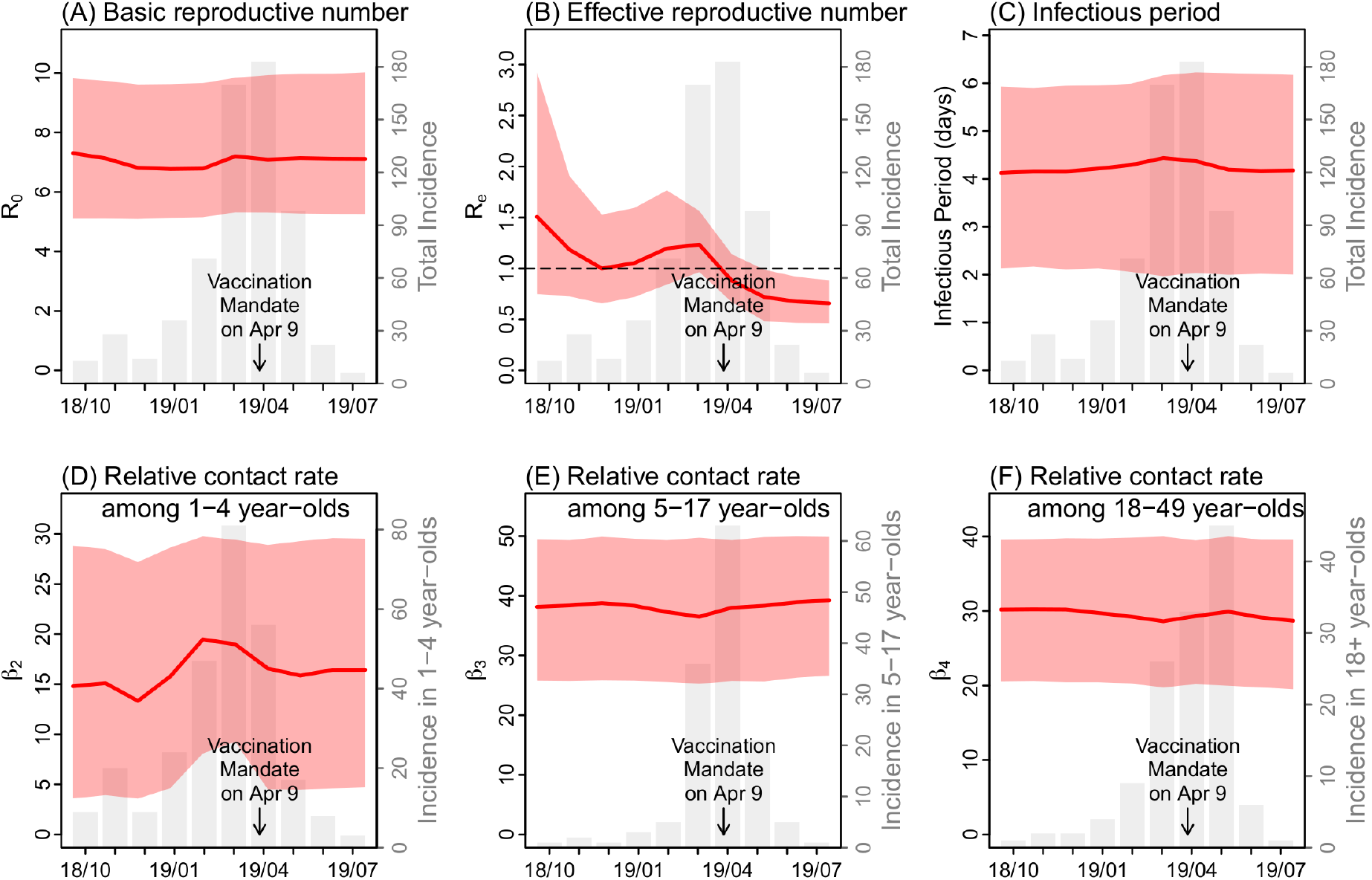
Estimates of key model parameters: (A) the basic reproductive number, (B) the effective reproductive number, (C) infectious period, (D) relative contact rate among 1-4 year-olds, (E) relative contact rate among 5-17 year-olds, and (F) relative contact rate among 18-49 year-olds. Red lines and surrounding areas (y-axis on the left) show the mean and 95% credible intervals of estimates pooled over all 10 model-filter runs (100,000 model realizations in total) made at the end of each month from Oct 2018 to July 2019. For comparison, the grey bars (y-axis on the left) show monthly incidence for all ages (A-C) or the related age groups (D-F).

The outbreak, however, took off again in early 2019 (Fig. 1). The estimated *R*_*e*_ increased and remained above 1 in the first three months of 2019 (Fig. 4B). In a perfectly mixed model, *R*_*e*_ is computed as the product of the basic reproductive number *R*_*0*_ and population susceptibility. In particular, the basic reproductive number *R*_*0*_ measures the transmissibility of an infection in a fully susceptible population; for measles, while often reported in the range of 12-18, *R*_*0*_ could vary from 1.4 to 770 (*6*). In this study, we estimated that *R*_*0*_ was approximately 7 during the entire outbreak (Fig. 4A). Population susceptibility, the other factor for *R*_*e*_, were to increase in a close population (i.e., without migration), could only do so slowly as infants lose maternal immunity. Thus, these two factors alone could not explain the sudden large increase in *R*_*e*_ (Fig. 4B). In this study, we utilized an age-structured model that enables more detailed analysis of the transmission dynamics. Indeed, our model-filter system detected an increase in contact rate among 1-4 year-olds during Jan-March 2019 (Fig. 4D); this increased contact rate along with the high susceptibility among 1-4 year-olds appeared to raise *R*_*e*_ above unity (Eqn 5 in Methods) and contribute to the re-surge of measles in early 2019. This finding was also consistent with reports of parents hosting “measles parties” to exposure unvaccinated children at the time (*7*). In contrast, estimated contact rates were relatively stable for other age groups (e.g., Fig. 4 E and F for 5-17 and 18-49 year-olds, respectively). As a result, infections increased quickly among 1-4 year-olds (Figures 1 and 4D), reaching a peak of around 80 cases in March. Meanwhile, infections also increased in other age groups including 5-17 and 18+ year-olds despite their low overall susceptibilities, due to interactions between age-groups (Fig. S2) and the high contact rates in these groups (Fig. 4 E and F).

The outbreak began to decline in Apr 2019, following more stringent public health interventions (*3, 4*). In particular, the model estimated that, thanks to extensive vaccination campaigns, susceptibility was reduced to 22.8% (95% CI: 19.3, 26.0%) in <1 year-olds, 4.3% (95% CI: 0.5, 8.9%) in 1-4 year-olds, and 2.4% (95% CI: 0.4, 4.4%) in 5-17 year-olds at the end of May. Consequently, the effective reproductive number *R*_*e*_ dropped below 1 from April onwards.

### Who acquired infection from whom?

Table 1 shows the estimated proportions of infections caused by each of the five age groups based on the estimated Who-Acquires-Infection-From-Whom (WAIFW) contact matrix (Eqn 2 in Methods). Children aged 1-4 years not only had the largest number of infections (42.8%), but also appeared to cause the largest number of infections in other age groups. Tallied over the entire study period (Oct 2018-July 2019), an estimated 51.6% (95% CI: 39.3, 63.1%) of the total cases were infected by 1-4 year-olds, compared to 25.2% (95% CI: 15.8, 35.5%) by 5-17 year-olds, 17.7% (95% CI: 10.4, 26.5%) by 18-49 year-olds, 4.5% (95% CI: 3.0, 6.4%) by <1 year-olds, and 1.0% (95% CI: 0.4, 1.8%) by 50+ year-olds. In particular, this age group caused around half of the infections in infants (44.6% or 45 cases) as well as the largest proportions of inter-group transmission to other age-groups (ranging from 12.9% to 5-17 year-olds to 40.1% to 50+ year-olds; Table 1).

**Table 1.** Estimated proportion of infections caused by each age-group. Rows show the receiving (i.e. infectee) age groups and columns show the source of infection (i.e. infector age group). The numbers are the mean (and 95% CI) estimates in percentage. For instance, for <1 year-olds (3^rd^ row), on average 16.3% of cases were infected by the same age group, 44.6% by 1-4 year-olds, 20.9% by 5-17 year-olds, 15.2% by 18-49 year-olds, and 3% by 50+ year-olds.

| Infectee age groups | Infector age groups |  |  |  |  |
| --- | --- | --- | --- | --- | --- |
|  | <1 year | 1-4 years | 5-17 years | 18-49 years | 50+ years |
| <1 year | 16.3 (12.3, 21.0) | 44.6 (35.5, 53.5) | 20.9 (13.9, 28.4) | 15.2 (9.6, 21.6) | 3.0 (1.4, 5.0) |
| 1-4 years | 1.8 (1.0, 3.0) | 85.8 (77.0, 91.9) | 7 (3.2, 12.9) | 5 (2.2, 9.3) | 0.3 (0.1, 0.6) |
| 5-17 years | 1.5 (0.9, 2.4) | 12.9 (6.9, 21.3) | 80.9 (70.4, 88.4) | 4.4 (2, 8) | 0.3 (0.1, 0.5) |
| 18-49 years | 2.4 (1.5, 3.7) | 18.6 (10.9, 28.0) | 9.1 (4.8, 14.9) | 69.5 (56.2, 80.8) | 0.4 (0.2, 0.8) |
| 50+ years | 15.5 (11.6, 19.9) | 40.1 (31.1, 49.3) | 19.7 (13, 27) | 15.6 (9.7, 22.4) | 9.1 (3.8, 16.3) |

### Estimated impact of vaccination campaigns

Figure 5 shows the estimated outbreak outcomes had there be no vaccination campaigns. Without the vaccination campaigns, the model estimated that the outbreak could continue to the end of 2019 and infect a total of 6196 (95% CI: 5, 8478) people by then or 6078 (95% CI: 5, 8433) during the observed outbreak period (i.e. Oct 2018 – July 2019), compared to 642 cases reported as of Aug 6, 2019. In addition, these infections would largely occur in infants under 1 and young children aged 1-4 years. During the observed outbreak period, there would be 1015 (95%: 0, 1430) infections in infants and 3052 (95% CI: 2, 4096) infections in 1-4 year-olds, more than 10 times of the reported numbers in these two age groups (i.e. 100 and 275, respectively, as of Aug 6, 2019; Table 2). Children aged 5-17 years would have the third largest number of infections, with 1107 (95% CI: 1, 1692) cases, 8 times of the reported number (i.e. 138 as of Aug 6, 2019).

**Table 2.**
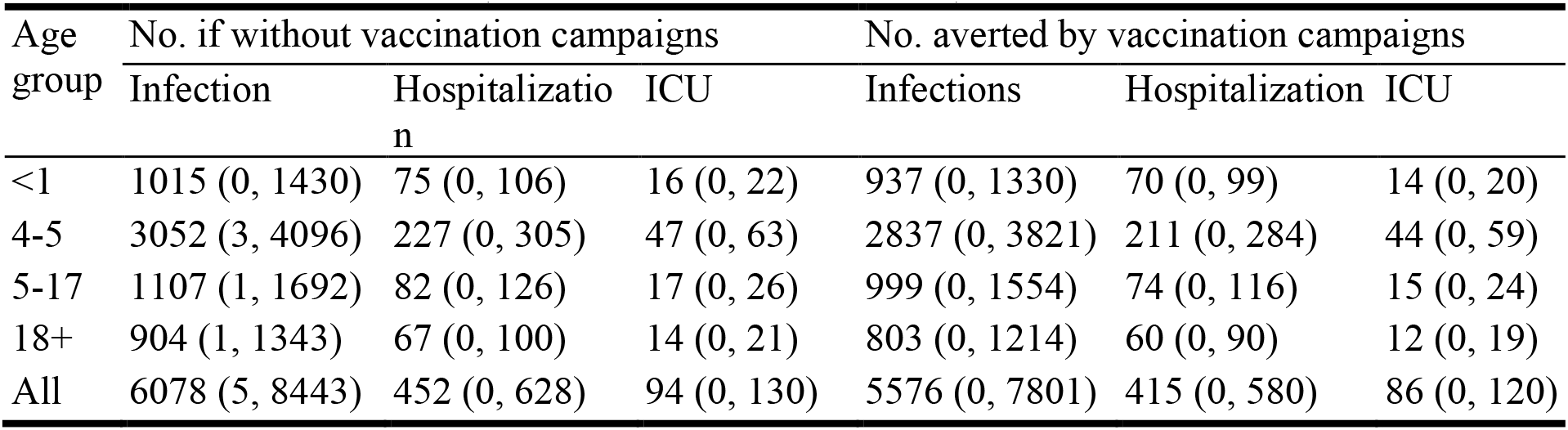
Estimated impact of vaccination campaigns during Oct 2018 – July 2019. Columns 2-4 show the total numbers of infections (2^nd^ column), hospitalizations (3^rd^ column), and individuals in intensive care unit (ICU) for different age groups (rows 3 to 6) and overall (last row), should there be no vaccination campaigns. Columns 5-7 show the number of infections, hospitalizations, and ICU cases averted by the vaccination campaigns, compared to data reported as of Aug 6, 2019. Numbers are the mean (and 95% confidence intervals) of 10,000 simulations.

**Fig. 5.**
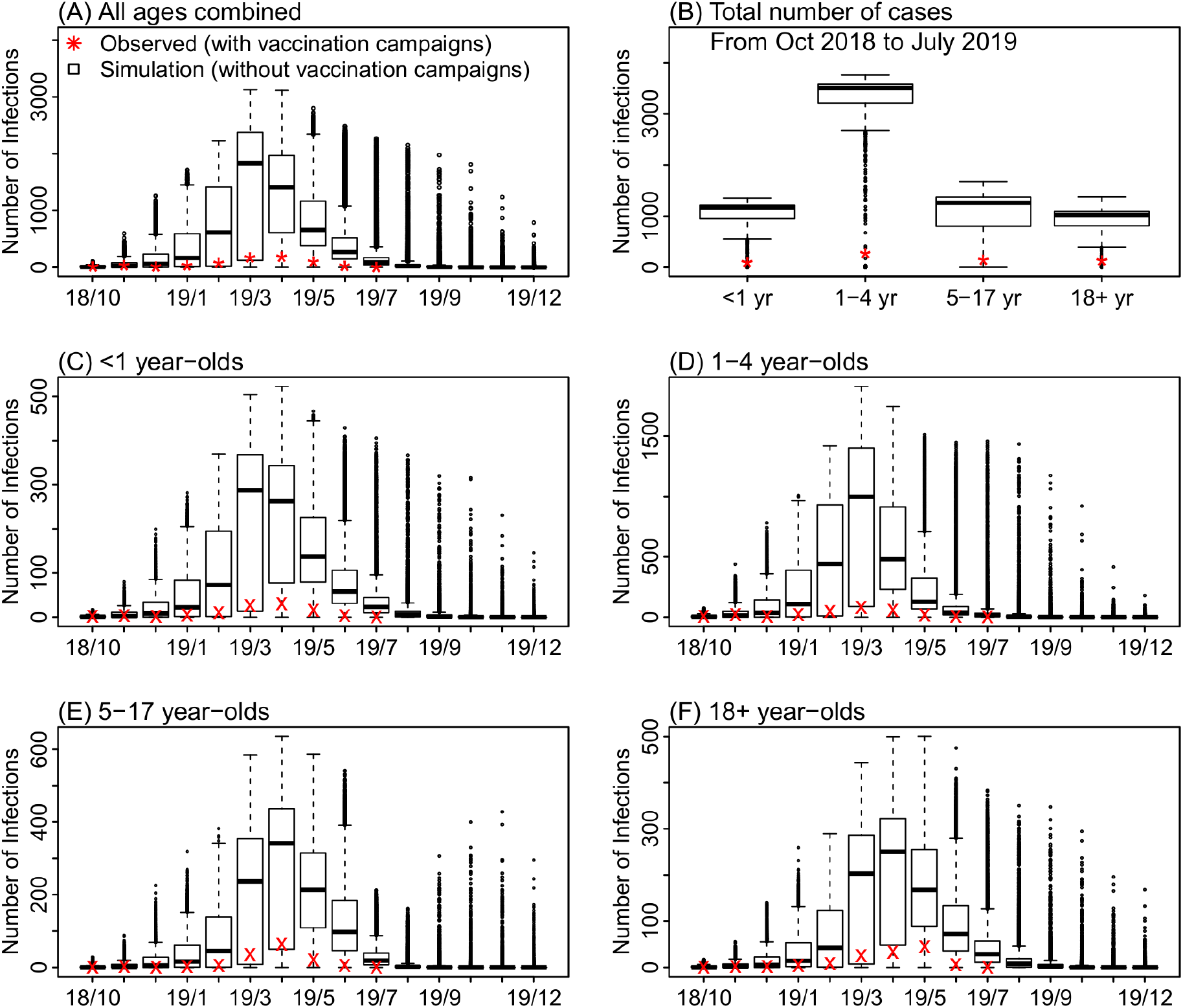
Estimated impact of vaccination campaigns. Box plots show simulated estimates of monthly incidence for all ages (A), percentage of cases reported in each age group (B), and monthly incidence for <1 year-olds (C), 1-4 year-olds (D), 5-17 year-olds (E), and 18+ year-olds (F), should there be no vaccination campaigns. Results are pooled over 10,000 model simulations. Horizontal thick lines show the median of model estimates; box edges show the 25^th^ and 75^th^ percentiles; whiskers show the 2.5^th^ and 97.5^th^ percentiles; and dots show outliers. For comparison, stars (*) in A and B show monthly incidence for all ages and the age distribution, reported as of Aug 6, 2019; crosses (x) in C-F show age-grouped monthly incidence estimated from health reports.

Measles can cause severe diseases. According to data by Apr 24, 2019, among 390 individuals with measles, 29 were hospitalized, of which six needed intensive care (*8*). Assuming the same ratios among all cases, without the vaccination campaigns, during the observed outbreak period there would be 452 (95% CI: 0, 628) hospitalizations, including 94 (95% CI: 0, 130) needing intensive care, and the majority would be in young children under 4 (Table 2).

## Discussion

Using a model-filter inference system, we have reconstructed in detail the transmission dynamics of the measles outbreak in an Orthodox Jewish community in NYC during Oct 2018 – July 2019. We have estimated the population characteristics (e.g. age-specific susceptibilities) and epidemiological parameters (e.g. reproductive numbers) as well as subtle changes in key parameters (e.g. contact rates) that are critical to the transmission of measles. Using model simulation and the posterior estimates from the model-inference system, we are also able to estimate the impact of vaccination campaigns implemented during the outbreak, including numbers of infections and hospitalizations averted, for each age group. These latter findings echo those from previous studies (*9-11*) and again highlight the severity of measles disease, should there be no effective infection and transmission controls (in particular, vaccination).

Our analyses estimate that around a quarter of young children aged 1-4 in the affected community were susceptible at the onset of the outbreak, likely due to delayed vaccination. Indeed, 94% (101/108) of the early infections in children were unvaccinated (*12*). In contrast, vaccination rate remained high in older children 5-17 years, with an estimated 94% immune to measles. This difference may be due to better compliance with vaccination regulation at school entry, or a result of vaccination campaigns in response to previous outbreaks (e.g., a large outbreak occurred in the same community in 2013 (*13*)). Nevertheless, the large number of unvaccinated children under 4 was sufficient to cause many infections in late 2018, predominantly in the same age group (Fig. 1 and references (*2, 12, 14*)). This observation highlights the importance of vaccination compliance with both MMR vaccine doses, especially given the long lag between the two vaccine doses. In addition, recommending the second MMR dose earlier than the currently scheduled age 4-6 years could allow pro-vaccine parents to fully vaccinate their children sooner and reduce the number of susceptible children overall.

Our study also reveals intricate interplays of population dynamics and measles transmission. While the high susceptibility in children under 4 was likely responsible for the early spread of measles, our estimates suggest that the second and more severe part of the outbreak in 2019 was likely due to increased infectious contact among this age group, likely facilitated by parents hosting “measles parties” that intentionally bring unvaccinated children together and expose them to those sick with measles (*7*). As shown in Fig. 4, the increase in infectious contact interacting with the high susceptibility in 1-4 year-olds was able to raise the effective reproductive number *R*_*e*_ to above unity—the threshold for an epidemic to occur—and aggravate the outbreak in 2019, despite earlier public health efforts that had reduced *R*_*e*_ to below 1 in late 2018. Similar disease-related gatherings have been noted in previous measles outbreaks (*15*) as well as other disease outbreaks (*16*). These activities create further challenges for the control of measles spread and stress the need for enhanced public health education.

In addition, the intensified measles outbreak not only affected children with delayed vaccination, but also a large number of infants under 1, who were too young to receive their first dose of MMR vaccine in the US. At least 100 infants under 1 were infected with measles during the 10-month outbreak period, despite extensive infection and transmission control efforts including vaccinating infants 6 months or older and post-exposure prophylaxis with immune globulin given to those under 6 months (*2, 14*). This was largely a result of the high susceptibility in infants. Our model-inference system estimates that about half of infants were susceptible by age 1 and that nearly half of the 100 infant cases were infected by 1-4 year-olds (Table 1). In addition, our simulations suggest that, without the vaccination campaigns, the number of infections in infants would be over 10 times higher than observed (Table 2). These findings demonstrate the rippling effects of vaccine hesitancy beyond the risk posted to age-eligible children with delayed vaccination. These findings also suggest that administration of the first dose of routine MMR vaccine earlier than the current 1 year age-limit in the US may be necessary to protect infants should high level of vaccine hesitancy persist. Of note, the World Health Organization (WHO) recommends administering the first dose of measles vaccine at 9-12 months of age for routine vaccination programs and as early as 6 months for settings such as during an outbreak (*17*).

Our model simulations, consistent with many previous studies (*11, 15, 18*), demonstrate the significant public health impact of vaccination in controlling measles outbreaks. Without the implemented vaccination campaigns, the severity of the measles outbreak—including number of infections, hospitalizations, and severe infections needing intensive care—would be about 10 times worse than observed (Table 2). These estimates, however, did not include the long-term health impacts on affected individuals, particularly young children (*10, 19, 20*), nor the enormous economic burdens (*13, 21, 22*).

We note several limitations of our study. First, we did not explicitly model the impact of public health interventions other than the vaccination campaigns, due to a lack of data. During the outbreak, such efforts included prescreening patients prior to presence for treatment, post-exposure prophylaxis, and closing schools out of vaccination compliance (*2, 12, 14, 23, 24*). Of note, however, here the estimated basic reproductive number *R*_*0*_ was around 7, lower than the 12-18 range based on epidemics in the pre-vaccine era (*25*); and the estimated infectious period was around 4 days (Fig. 4C), at the lower end of the commonly used range of 4-6 days (*26*). The lower *R*_*0*_ and shorter infectious period could be a result of the aforementioned public health interventions. Second, there were uncertainties in the accuracy of case reporting. Because the incidence data used here were published on Aug 6, 2016, a later revision of case reports, we used a relatively high but broad prior range for the reporting rate (i.e. 80-100%). In addition, our model-inference system explicitly accounted for observational errors (Eqn 7a and 7b). Third, due to a lack of contact data and for simplicity, we set all terms related to group-1 (i.e. <1 year-olds) in the WAIFW matrix to the same as the contact rate within the group (Eqn 2). The model formulation may have led to under-estimation of the proportions of infection in <1 year-olds attributable to the same age group and/or 1-4 year-olds, given the likely more frequent contact within the same age group and with similar ages (i.e. 1-4 years) due to more shared settings such as daycares and pediatric hospitals. Lastly, there were uncertainties regarding the settings of vaccination campaigns. Nevertheless, sensitivity analysis showed that our main estimates were robust to a wide range of assumptions (Table S2).

In summary, using a comprehensive model-inference system, we have reconstructed transmission dynamics of the recent measles outbreak in NYC in great detail. Our estimates highlight the importance of vaccination in protecting children as well as public health education to reduce activities that unnecessarily expose children to risk of measles infection. Further, in light of the persistent vaccine hesitancy, revising current vaccination recommendations may allow pro-vaccine parents to vaccinate and protect their children more effectively.

## Materials and Methods

The measles outbreak occurred predominantly among members of the Orthodox Jewish community in Williamsburg and Borough Park, two neighborhoods in located in Brooklyn, NYC. As such, we focused on modeling the outbreak in this subpopulation. Estimated based on the Jewish Community Study of New York (*27*), approximately *N*=165,970 Orthodox Jews live in these two NYC neighborhoods, of which 4552 (2.7%), 18,208 (11%), 59,176 (35.7%), 60,445 (36.4%), and 23,589 (14.2%) are <1, 1-4, 5-17, 18-49, and 50+ years, respectively.

### Estimating monthly incidence by age group

Monthly measles incidence aggregated over all ages from Sep 2018 to Aug 2019 and the age distribution of case-patients over the entire outbreak were published on the NYC DOHMH website (*3*). Of note, 1 case was reported in Sep 2018 (i.e., the initial case) and none were reported in Aug 2019 at the time of this writing. In addition, the numbers of reported cases were subsequently revised by the DOHMH (often adjusted upwards, presumably from retrospective case identification) and, as such, varied over time. To estimate the monthly incidence for each age group, we used the age distribution of cases reported in earlier health reports/alerts (*5*) to apportion the total incidence for each month. For months without age information, we used estimates either from the preceding month or the following month back-calculated from the overall age distribution.

### Transmission model

The transmission model used here was similar to described in our previous study (*28*). As illustrated in Fig. S1, the model represents the susceptible-exposed-infectious-recovered (SEIR) disease dynamics with five age groups (i.e. <1, 1-4, 5-17, 18-49, and 50+ years) to account for population differences by age group (e.g., susceptibility and contact rate), routine two-dose vaccination at ages 1 and 5, and immunization during the vaccination campaigns per Eqn 1:

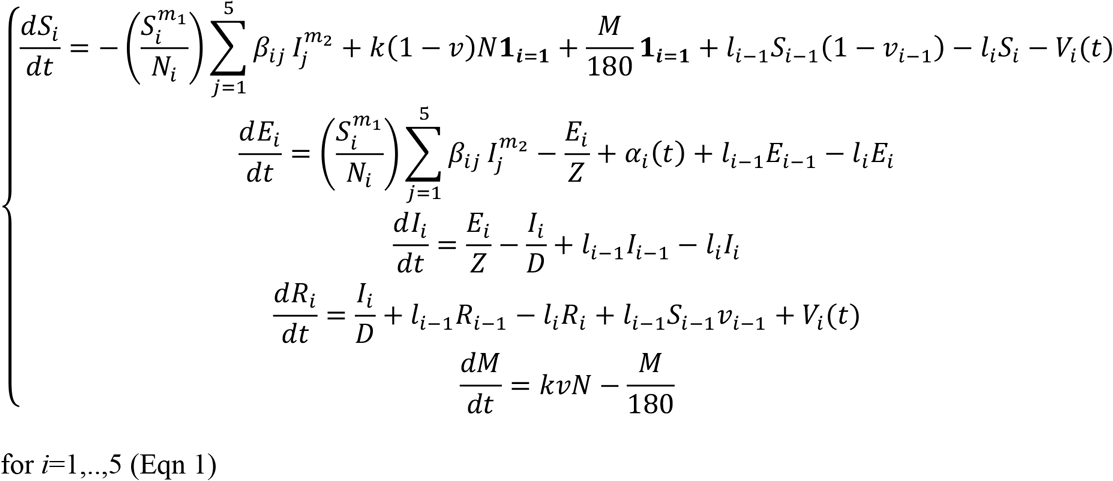

where *S*_*i*_, *E*_*i*_, *I*_*i*_, *R*_*i*_ and *N*_*i*_ are, respectively, the numbers of susceptible, exposed (i.e. latently infected), infectious, recovered (and/or immunized) people and population size in the *i*-th age group; *M* is the number of infants with maternal immunity, which decays exponentially with a mean duration of 180 days; *t* is time in days. *V*_*i*_(*t*) is the number of people in group-*i* immunized by the vaccination campaigns on day-*t* (described in detail in the next section). The exponents *m*_*1*_ and *m*_*2*_ describe the level of inhomogeneous mixing (*29, 30*); and *m*_*1*_=*m*_*2*_=1 represents homogeneous mixing. *Z* and *D* are the latent and infectious period, respectively.

To model the different contact rates within and between age groups, we used 7 parameters for the Who-Acquires-Infection-From-Whom (WAIFW) matrix as follows:

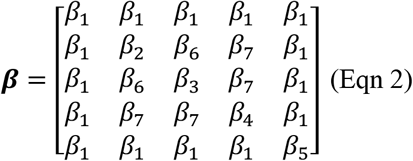

where *β*_*1*_ *to β*_*5*_ represent within-group contact for the five age groups and *β*_*6*_ and *β*_*7*_ represent mixing between siblings and child-parent, respectively. For simplicity, we set all interactions with group 1 (i.e. <1 year) or group 5 (50+ years) to *β*_*1*_, the lowest contact rate. For group-3 (5-17 years), to capture the varying contact rate following school schedules, we adjusted *β*_*3*_ for each date per the school calendar in NYC as:

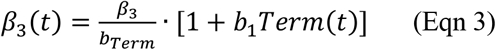

where *b*_*1*_ is the amplitude of school term-time forcing; *Term*(*t*) is set to 1 for school days and −1 for non-school days; and *b*_*Term*_ is the yearly average of 1+*b*_*1*_*Term*(*t*) (*31*).

The basic reproductive number *R*_*0*_, defined as the average number of secondary infections caused by a primary case-patient in a *naïve* population, reflects the transmissibility of an infection. In an age-structured model, *R*_*0*_ is computed as:

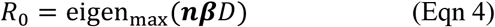

where eigen_max_(·) denotes the function giving the largest eigenvalue of a matrix, and ***n*** is a diagonal matrix with elements *n*_*i*_=*N*_*i*_/∑*N*_*i*_ (*i*=1, …, 5 here), i.e. the fraction of population in group-*i*. Based on this relationship between *R*_*0*_ and the ***β*** matrix, we reparametrized the model to include *R*_*0*_ as a model parameter by setting *β*_*1*_ to 1 and estimating the relative magnitude of *β*_*2*_–*β*_*6*_, all scaled to *R*_*0*_. In the current mass vaccination era, most people are immune via vaccination. To reflect the potential of an infection to cause an epidemic in a partially susceptible population, the effective reproductive number, *R*_*e*_, accounts for population susceptibility and is computed as:

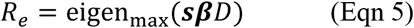

where ***s*** is a diagonal matrix with elements *s*_*i*_=*S*_*i*_/*N*_*i*_ (*i*=1, …, 5 here), i.e. the susceptibility in group-*i*.

To model the demographic processes, *k* is the birth rate (2.7 per 1000 person-year here (*27*)); *v* is the immunity level in mothers, approximated by the susceptibility of the child-bearing age-group (i.e., 18-49 year-olds); *N* is total population size; and **1**_i=1_ is an indicator function, with value 1 for group-1 (<1 year-olds) and 0 for all other groups. Thus, the term *k*(1-*v*) **1**_i=1_ (1^st^ line in Eqn 1) is the number of susceptible newborns and *kvN* (last line in Eqn 1) is the number of newborns with maternal immunity. The term *l*_*i*_ is the rate of aging for group-*i* (i.e. the inverse of the sojourn time in each age group) with *l*_*0*_ set to 0 and *l*_*5*_ set to the death rate. The term *v*_*i*_ (for *i*=1 and 2) is the vaccination rate for the two doses of vaccine (i.e., *v*_*i*_=0 for *i*=0, 3, and 4). In this study, we set *v*_*1*_ (1^st^ dose) to 0.9 times the immunity level of group-2 (i.e., assuming a 90% vaccine efficacy) and *v*_*2*_ (2^nd^ dose) to 0.7 for days before May 2019; for days afterwards, we used 0.8 for *v*_*1*_ and 0.9 for *v*_*2*_, corresponding to a 1-(1-0.8)(1-0.9)=98% overall vaccination rate.

Based on NYC health reports/alerts (*2, 12, 14*), we seeded the model, via the parameter *α*_*i*_(*t*), with 3 cases in group-2 (i.e. 1-4 year-olds)—one each with rash onset on Sep 30, Oct 15, and Oct 30, 2018, respectively—and 1 case each in group-3 (5-17 year-olds) and group-4 (18-49 year-olds), both during the winter recess (from 12/24/18 to 1/1/19).

### Modeling the vaccination campaigns

To contain the outbreak, the NYC DOHMH conducted extensive vaccination campaigns and administered 31,790 doses of MMR vaccine to children under 19 years in Williamsburg and Borough Park by July 2019 (*3*). However, information on the age and immune status of vaccinees was not reported. In this study, per the health reports/alters (*3, 12*), we assumed there were two phases of vaccination campaign: 1) Oct 2018 – Feb 2019, during which 7000 children, 90% Orthodox Jewish (15% were <1 year, 65% 1-4 years, and 20% 5-17 years) were vaccinated; and 2) Mar – July 2019, during which 24,790 children, 60% Orthodox Jewish (10%, 40%, and 50%, respectively, were <1, 1-4, and 5-17 years) were vaccinated. For reference, Orthodox Jews made up for approximately 30% of the total population in the two affected neighborhoods.

For <1 year-olds, immunization could fail due to residual maternal immunity; thus, we assumed a 85% immunization success rate for group-1. For those above 1 year, some vaccinees might have received 1 or 2 doses of vaccine and higher the population susceptibility, the less likely a vaccinee had been immune before the additional vaccine dose. As such, we assumed the immunization success rate was twice the group-specific susceptibility for 1-4 year-olds and three times that for 5-17 year-olds or at a minimum of 25% and a maximum of 75%. We further assumed a 10-day delay in vaccine effect and computed the daily number of individuals vaccinated per a gamma distribution (mean=30 days and standard deviation=21 days for Phase-1 and mean=56 and standard deviation=15 days for Phase-2 such that it peaked ∼1 week after Apr 9, 2019 when the city implemented a vaccination mandate). The estimated numbers matched with the reports (e.g., 1740 doses by our model vs ∼1600 doses given to children under 5 as reported (*32*) and 1142 doses by our model vs ∼1000 doses given in March 2019 as reported (*7, 33*)). These daily numbers were then included in the transmission model [i.e. *V*_*i*_(*t*) in Eqn 1]. Sensitivity to model assumptions were tested as described below.

### Estimation of model state variables and parameters

To estimate the model state variables (i.e., *S*_*i*_, *E*_*i*_, *I*_*i*_, *R*_*i*_, and *M*) and parameters (*β*_*2*_ *to β*_*7*_, *R*_*0*_, *b*_*1*_, *D, Z, m*_*1*_, *and m*_*2*_), we fitted the model to the reported monthly overall incidence and the estimated monthly age-grouped incidence using a particle filter (*34*). Briefly, we first initialized a suite of model realizations (termed “particles”, *N*=10,000 here) using Latin Hypercube sampling (*35*) from the prior distribution of state variables and parameters (Table S1). The particle filter then sequentially incorporated the monthly incidence to the model via repeated prediction-update cycles. In each cycle (i.e., each month here), the particles were stochastically integrated forward in time for a month per the model (i.e., Eqn 1; this generates the prediction). To update the model state including all model variables and parameters, at the end of each month, the model-estimated incidence was aggregated for the month, adjusted by the reporting rate for that month (estimated simultaneously by the filter), and used to compute the likelihood of each particle (described below). The posterior of model state was then computed using Bayes’ rule (*34, 36*) and the particles resampled and updated—those with high posterior probabilities were retained and those with very low posterior probabilities discarded.

To allow for a wider observational variance than, e.g., the Poisson process, we heuristically modeled the observations using a multivariate Gaussian distribution (i.e. the likelihood function):

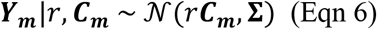

where ***Y***_***m***_ is the vector of monthly incidence reported for month-*m*, including the monthly incidence for individual age-groups (i.e., <1, 1-4, 5-17 and 18+ years; note that 18-49 and 50+ year-olds were combined due to a lack of data for these two groups separately) and all ages combined. Correspondingly, ***C***_***m***_ is the vector of monthly incidence estimated by the model; and *r* is the reporting rate and, for simplicity, assumed the same for all ages. **∑** is the covariance matrix, with the off-diagonal terms set to 0. To account for uncertainties in the estimated age-grouped incidence, the variance (∑_*ii*_, *i* = 1, …, 4) for each of the 4 aforementioned age-groups was heuristically computed as:

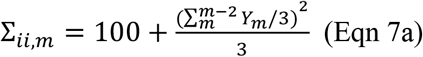

That is, the observational variance is proportional to the average incidence in the preceding two months (if available) and the current month, plus a baseline constant. For the overall incidence with detailed data, a smaller variance was used:

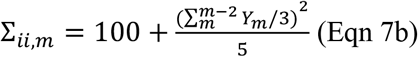

As there were great uncertainties in the susceptibilities of the younger age groups, we tested prior ranges from 10-40% for 1-4 year-olds, 5-20% for 5-17 year-olds, and 5-15% for 18-49 year-olds. For the basic reproductive number *R*_*0*_, we tested prior values ranging from 5 to 12 (note these values were lower than the oft-reported 12-18 range (*6, 25*)). To optimize the model-filter system, we parsed these wide ranges into smaller segments and tested all combinations by permutation (5040 in total; see specific prior ranges in Table S1). To account for model stochasticity, we ran the model-filter system 5 times for each prior combination and 10 times for the final prior select. We then selected the optimal priors based on the model-goodness-of-fit to the data (minimal root-mean-square-error and maximal correlation and likelihood) over the period of Oct 2018 – July 2019 as well as accuracy of the one-step-ahead predictions (recall that the particle filtering process comprises sequential prediction-update cycles) for the period of Oct 2018 – March 2019 (i.e. before the emergency vaccination mandate). We pooled all 10 final runs (10,000 particles each run and 100,000 model realizations in total) to compute the posterior estimates (e.g., mean and 95% CI).

### Sensitivity analysis on vaccination campaigns settings

To test the sensitivity of model results to assumptions on vaccination campaign settings, we tested the model-filter system using the following alternative scenarios:

1. For the 2^nd^ phase (Mar-July 2019), 90% (vs. 60% in the baseline scenario) of the vaccine doses were given to members of the Orthodox Jewish community;
2. For the 2^nd^ phase, the age distribution of vaccinees was the same as the 1^st^ phase (i.e. 15%, 65%, and 20%, respectively, for <1, 1-4, and 5-17 year-olds vs. 10%, 40%, and 50% for the three groups in the baseline scenario);
3. For the 2^nd^ phase, 90% of the vaccine doses were given to Orthodox Jewish and the age distribution of vaccinees was the same as the 1^st^ phase (i.e. 15%, 65%, and 20% for <1, 1-4, and 5-17 year-olds, respectively).

Because population susceptibility would be affected by the number of individuals immunized by the vaccination campaigns, we tested susceptibility ranges 10-40% for 1-4 year-olds, 5-20% for 5-17 year-olds, and 5-15% for 18-49 year-olds, divided in to small segments as for the baseline scenario (140 different combinations for each alternative scenario; Table S1). For simplicity, we used the same optimal prior ranges for the parameters under the baseline scenario in this sensitivity analysis.

### Evaluating the impact of vaccination campaigns

To estimate the impact of vaccination campaigns, we generated model-simulated counterfactuals—i.e., outbreak outcomes should there be no vaccination campaigns implemented—using the posterior mean estimates of group-specific initial population susceptibilities and model parameters for each month. We ran the model (10,000 realizations) stochastically up to the end of 2019 to test how long the outbreak could last without intervention; for months after Aug 2019, parameters estimated at the end of July 2019 were used.

## Data Availability

All related data are publicly available at the NYC DOHMH website (https://www1.nyc.gov/site/doh/health/health-topics/measles.page).

## Acknowledgements

This study was supported by the National Institute of Allergy and Infectious Diseases (1R01AI145883-01). I thank all public health personnel and individuals involved in combating the measles outbreak and New York City Department of Health and Mental Hygiene for making the incidence data publicly available. I also thank Columbia University Mailman School of Public Health for access to high performance computing.

## Conflict of interest

None.

## Supplementary Materials

**Table S1.** Main model parameters and prior ranges tested. In total, we tested 5040 combinations of prior ranges. Each combination was used as the lower and upper bounds of Latin Hypercube sampling. The optimal prior used in the final model-filter runs are bolded if multiple ranges were tested.

| Parameter | Symbol/Equation | Ranges tested |
| --- | --- | --- |
| Initial susceptibility in <1 year-olds | $S_1(t=0)$; Eqn 1 | Based on susceptibility in 18-49 year-olds (i.e., the mothers) |
| Initial susceptibility in 1-4 year-olds | $S_2(t=0)$; Eqn 1 | [5, 15], [10, 20], [15, 25], **[20, 30]**, [25, 35], [30, 40], [35, 45]% |
| Initial susceptibility in 5-17 year-olds | $S_3(t=0)$; Eqn 1 | **[4, 8]**, [5, 10], [5, 15], [10, 20], [15, 25]% |
| Initial susceptibility in 18-49 year-olds | $S_4(t=0)$; Eqn 1 | **[4, 8]**, [5, 10], [5, 15], [10, 20]% |
| Initial susceptibility in 50+ year-olds | $S_5(t=0)$; Eqn 1 | [4, 8]% |
| Initial number of infants with maternal immunity | $M(t=0)$; Eqn 1 | Based on susceptibility in 18-49 year-olds (i.e., the mothers) |
| Latent period | $Z$; Eqn 1 | [7, 9] days |
| Infectious period | $D$; Eqn 1 | [2, 6] days |
| Mixing parameter for the susceptibles | $m_1$; Eqn 1 | **1** (perfect mixing), [0.95, 1], [0.9, 0.95] |
| Mixing parameter for the infectious | $m_2$; Eqn 1 | **1** (perfect mixing), [0.95, 1], [0.9, 0.95] |
| Relative contact rate among <1 year-olds | $\beta_1$; Eqn 2 | Set to 1 |
| Relative contact rate among 1-4 year-olds | $\beta_2$; Eqn 2 | [3, 30] |
| Relative contact rate among 5-17 year-olds | $\beta_3$; Eqn 2 | [25, 50] |
| Relative contact rate among 18-49 year-olds | $\beta_4$; Eqn 2 | [20, 40] |
| Relative contact rate among 50+ year-olds | $\beta_5$; Eqn 2 | [1, 5] |
| Relative contact rate between 1-4 and 5-17 year-olds (sibling interactions) | $\beta_6$; Eqn 2 | [1, 5] |
| Contact rate between 18-49 and 1-4 or 5-17 year-olds (parent-child interactions) | $\beta_7$; Eqn 2 | [1, 5] |
| Amplitude of school term-time forcing | $b_1$; Eqn 3 | [0.25, 0.75], **[0.5, 1]** |
| Basic reproductive number | $R_0$; Eqn 4 | **[5, 10]**, [7, 12] |
| Reporting rate | $r$; Eqn 6 | [80, 100]% |

**Table S2.** Comparison of model estimates on initial susceptibility and model performance under different assumptions on vaccination campaigns. The baseline setting is as reported in the main text and alternative settings 1 to 3 are as described in the section “*Sensitivity analysis on vaccination campaigns settings*.” The results are summarized by pooling all 10 model-filter runs (10,000 particles each run and 100,000 model realizations in total). The numbers are the mean and, for the susceptibilities, 95% credible intervals in the parentheses. The initial susceptibilities, estimated at the end of Sep 2018, were computed by adding the total numbers of individuals immunized by the vaccination campaigns in Oct 2018 to the posterior estimates at the end of Oct 2018.

|  | Age group | Model Settings on Vaccination Campaigns |  |  |  |
| --- | --- | --- | --- | --- | --- |
|  |  | Baseline | Alternative 1 | Alternative 2 | Alternative 3 |
| Estimated initial susceptibility (%) at end of Sep 2018 | <1 year | 53.2 (49, 57.5) | 54.2 (50, 58.4) | 54.2 (50, 58.5) | 54.2 (50, 58.5) |
|  | 1-4 years | 24.9 (20.4, 29.7) | 24.9 (20.4, 29.7) | 24.9 (20.4, 29.7) | 29.9 (25.4, 34.7) |
|  | 5-17 years | 6.0 (4.1, 7.9) | 7.5 (5.1, 9.9) | 7.5 (5.1, 9.9) | 6.0 (4.1, 7.9) |
|  | 18-49 years | 6.0 (4.1, 7.9) | 7.5 (5.2, 9.9) | 7.5 (5.2, 9.9) | 7.5 (5.2, 9.9) |
|  | 50+ years | 6.0 (4.1, 7.9) | 6.0 (4.1, 7.9) | 6.0 (4.1, 7.9) | 6.0 (4.1, 7.9) |
| Log-likelihood |  | -255.25 | -265.52 | -255.25 | -257.42 |
| Relative error of total number of cases over the outbreak | <1 year | 0.39% | -11.96% | -11.56% | -17.91% |
|  | 1-4 years | -0.04% | -7.80% | -6.90% | -8.07% |
|  | 5-17 years | -4.13% | 14.38% | 0.33% | 1.84% |
|  | 18+ years | -4.70% | 7.24% | 5.97% | 7.95% |
|  | All ages | -1.79% | -0.65% | -3.48% | -4.25% |
| Root-mean-square-error (RMSE), over Oct 2018 – July 2019 | <1 year | 1.74 | 1.86 | 1.80 | 2.55 |
|  | 1-4 years | 8.72 | 8.13 | 8.17 | 7.70 |
|  | 5-17 years | 3.91 | 5.27 | 3.00 | 5.10 |
|  | 18+ years | 5.08 | 4.62 | 4.30 | 4.25 |
|  | All ages | 7.31 | 7.52 | 5.88 | 6.29 |
| Correlation, over Oct 2018 – July 2019 | <1 year | 0.99 | 1.00 | 1.00 | 0.99 |
|  | 1-4 years | 0.95 | 0.96 | 0.96 | 0.97 |
|  | 5-17 years | 0.99 | 0.97 | 0.99 | 0.97 |
|  | 18+ years | 0.97 | 0.97 | 0.97 | 0.97 |
|  | All ages | 0.99 | 0.99 | 1.00 | 1.00 |
| 1-step-head prediction RMSE, over Oct 2018 – Mar 2019 | <1 year | 4.37 | 4.46 | 4.40 | 4.60 |
|  | 1-4 years | 17.13 | 17.34 | 17.35 | 25.12 |
|  | 5-17 years | 8.27 | 8.39 | 8.37 | 8.50 |
|  | 18+ years | 4.34 | 4.93 | 4.93 | 5.06 |
|  | All ages | 27.79 | 28.23 | 28.16 | 35.02 |

**Fig. S1.**
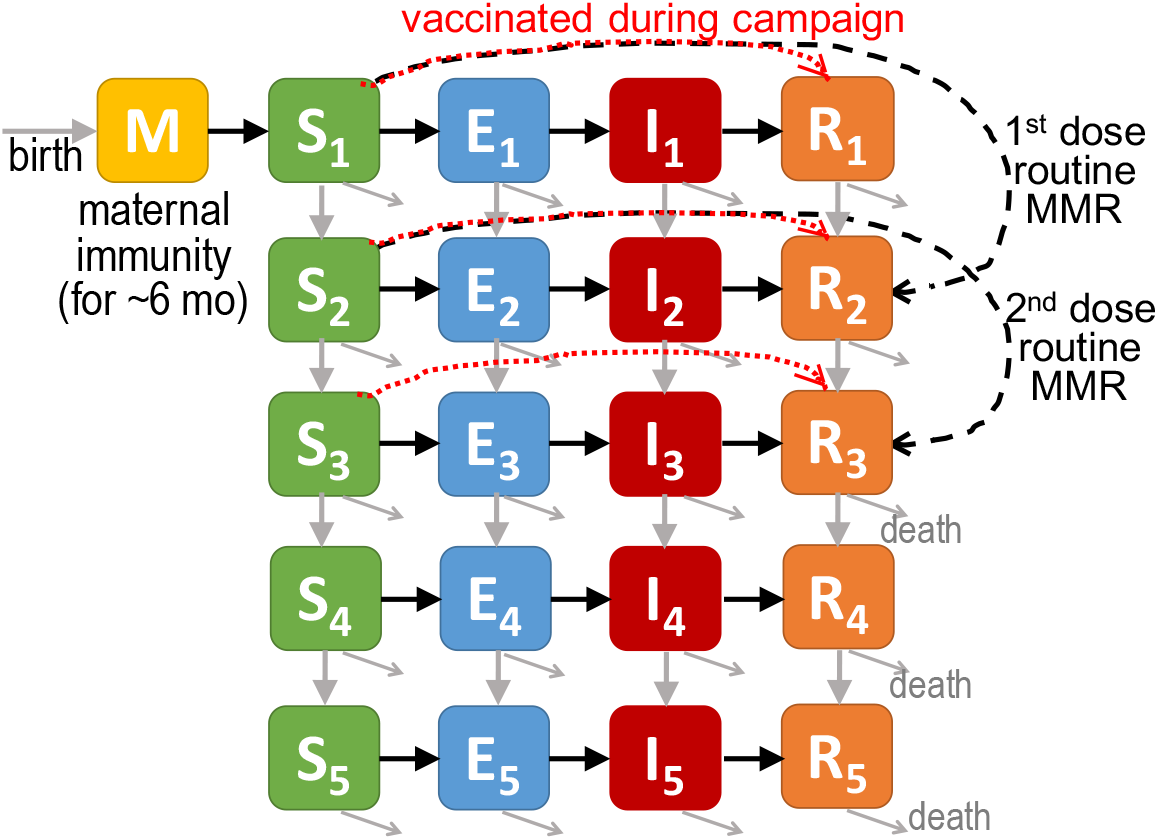
Schematic of the measles transmission model. Measles transmission model follows the susceptible (S), exposed (E) and latently infected, infectious (I), and recovered (R) and/or immunized SEIR dynamics and includes 5 age-groups as indicated by the subscripts (i.e., <1, 1-4, 5-17, 18-49, and 50+ year-olds, respectively) and a group (M) for infants with maternal immunity. Black solid arrows show the disease-related processes; grey solid arrows show the demographic processes including birth (horizontal), aging (vertical), and death (tilted). Black dashed arrows show processes related to the routine 2-dose measles vaccination where susceptible individuals are vaccinated at ages 1 and 5 and move to the respective immune groups. Red dotted arrows show processes related to vaccination of susceptible individuals under 18 during the vaccination campaigns.

**Fig. S2.**
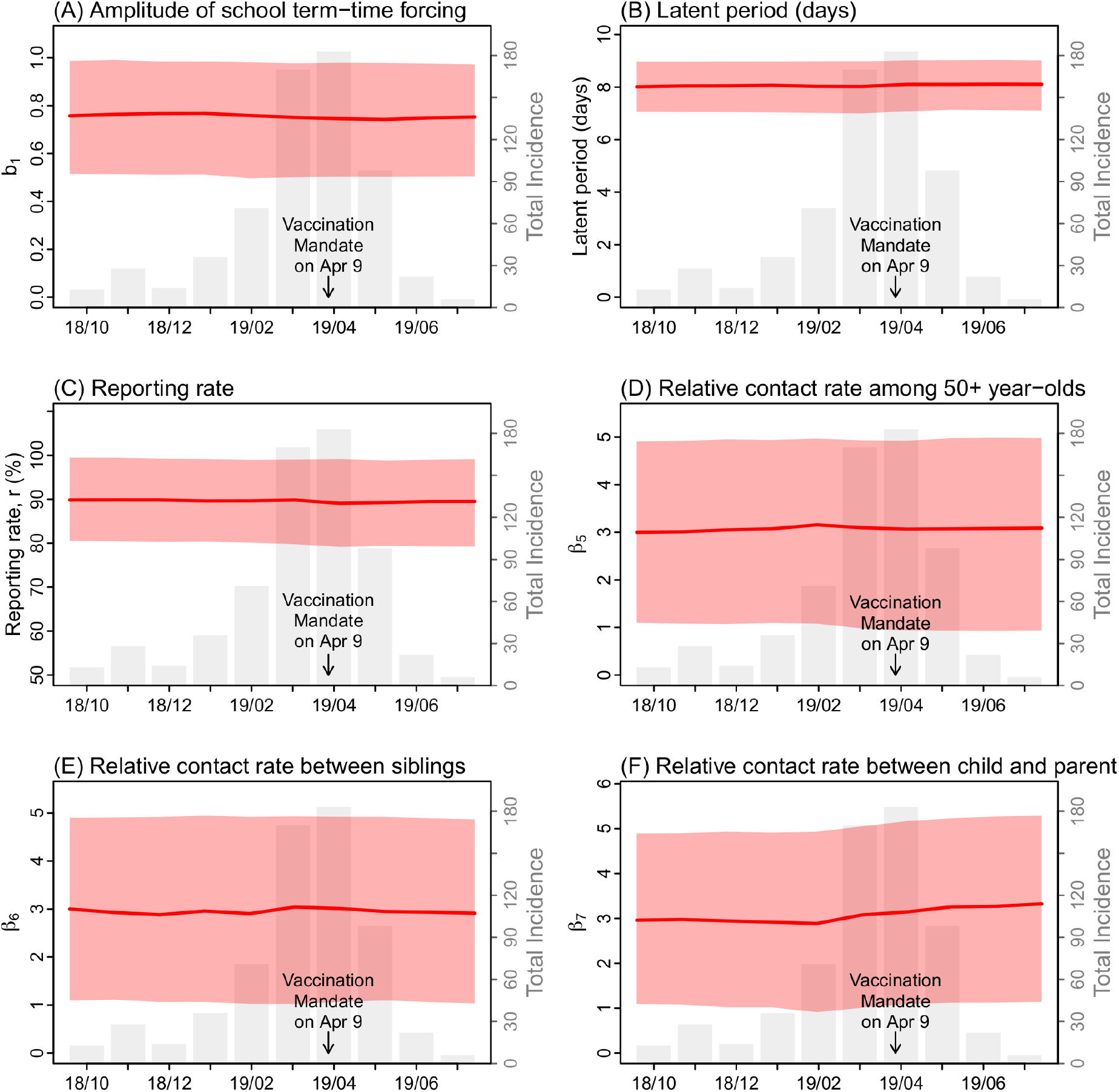
Estimates of model parameters not listed in Fig. 4: (A) amplitude of school term-time forcing, (B) latent period, (C) reporting rate, (D) relative contact rate among 50+ year-olds, (E) relative contact rate between 1-4 and 5-17 year-olds (i.e. sibling interactions), and (F) relative contact rate between 18-49 and 1-4 or 5-17 year-olds (i.e. parent-child interactions). Red lines and surrounding areas (y-axis on the left) show the mean and 95% credible intervals of estimates pooled over all 10 model-filter runs (100,000 model realizations in total) made at the end of each month from Oct 2018 to July 2019. For comparison, the grey bars (y-axis on the left) show monthly incidence for all ages. Note that *m*_*1*_ and *m*_*2*_ are not shown as both optimal priors are the value 1 (Table S1).

## Notes

### Competing Interest Statement

The authors have declared no competing interest.

### Clinical Trial

N/A

### Author Declarations

All relevant ethical guidelines have been followed and any necessary IRB and/or ethics committee approvals have been obtained.

Any clinical trials involved have been registered with an ICMJE-approved registry such as ClinicalTrials.gov and the trial ID is included in the manuscript.

## References

1. V. K. Phadke, R. A. Bednarczyk, D. A. Salmon, S. B. Omer, Association Between Vaccine Refusal and Vaccine-Preventable Diseases in the United States A Review of Measles and Pertussis. Jama-J Am Med Assoc 315, 1149–1158 (2016).

2. New York City Department of Health and Mental Hygiene, ALERT # 39: Update on Measles Outbreak in New York City in the Orthodox Jewish Community. (2018). https://www1.nyc.gov/assets/doh/downloads/pdf/han/alert/2018/alert39-measles-outbreak.pdf.

3. New York City Department of Health and Mental Hygiene, Measles. (2019). https://www1.nyc.gov/site/doh/health/health-topics/measles.page.

4. New York City Department of Health and Mental Hygiene, Order of the Commissioner. (2019). 4/9/19. https://www1.nyc.gov/assets/doh/downloads/pdf/press/2019/emergency-orders-measles.pdf.

5. New York City Department of Health and Mental Hygiene, Health Alert Network. (2019). https://www1.nyc.gov/site/doh/providers/resources/health-alert-network.page.

6. F. M. Guerra, S. Bolotin, G. Lim, J. Heffernan, S. L. Deeks, Y. Li, N. S. Crowcroft, The basic reproduction number (R0) of measles: a systematic review. Lancet Infect Dis 17, e420–e428 (2017).

7. S. Scutti, New York City declares a public health emergency amid Brooklyn measles outbreak. (2019). 4/9/19. https://www.cnn.com/2019/04/09/health/measles-new-york-emergency-bn/index.html.

8. New York City Department of Health and Mental Hygiene, ALERT # 9: Citywide Recommendations during the Ongoing Measles Outbreak in New York City. (2019). https://www1.nyc.gov/assets/doh/downloads/pdf/han/alert/2019/recommendations-during-measles-outbreak.pdf.

9. M. J. Mina, B. T. Grenfell, C. J. E. Metcalf, Response to Comment on “Long-term measles-induced immunomodulation increases overall childhood infectious disease mortality”. Science 365, (2019).

10. M. J. Mina, C. J. E. Metcalf, R. L. de Swart, A. D. M. E. Osterhaus, B. T. Grenfell, Long-term measles-induced immunomodulation increases overall childhood infectious disease mortality. Science 348, 694–699 (2015).

11. P. A. Gastanaduy, S. Funk, P. Paul, L. Tatham, N. Fisher, J. Budd, B. Fowler, S. de Fijter, M. DiOrio, G. S. Wallace, B. Grenfell, Impact of Public Health Responses During a Measles Outbreak in an Amish Community in Ohio: Modeling the Dynamics of Transmission. Am J Epidemiol 187, 2002–2010 (2018).

12. New York City Department of Health and Mental Hygiene, ALERT # 2: Update on Measles Outbreak in New York City in the Orthodox Jewish Community. (2019). https://www1.nyc.gov/assets/doh/downloads/pdf/han/alert/2019/update-on-measles-outbreak-in-nyc-in-the-orthodox-jewish-community.pdf.

13. J. B. Rosen, R. J. Arciuolo, A. M. Khawja, J. Fu, F. R. Giancotti, J. R. Zucker, Public Health Consequences of a 2013 Measles Outbreak in New York City. JAMA Pediatr 172, 811–817 (2018).

14. New York City Department of Health and Mental Hygiene, ALERT # 38: Measles Outbreak in New York City in the Orthodox Jewish Community. (2018). https://www1.nyc.gov/assets/doh/downloads/pdf/han/alert/2018/alert38-measles-outbreak.pdf.

15. P. A. Gastanaduy, E. Banerjee, C. DeBolt, P. Bravo-Alcantara, S. A. Samad, D. Pastor, P. A. Rota, M. Patel, N. S. Crowcroft, D. N. Durrheim, Public health responses during measles outbreaks in elimination settings: Strategies and challenges. Human vaccines & immunotherapeutics 14, 2222–2238 (2018).

16. Centers for Disease Control and Prevention, Chickenpox (Varicella): Transmssion. (2018). December 31, 2018. https://www.cdc.gov/chickenpox/about/transmission.html.

17. W. H. Organization, Measles vaccines: WHO position paper, April 2017– Recommendations. Vaccine, (2017).

18. J. Lessler, C. J. E. Metcalf, F. T. Cutts, B. T. Grenfell, Impact on Epidemic Measles of Vaccination Campaigns Triggered by Disease Outbreaks or Serosurveys: A Modeling Study. Plos Medicine 13, (2016).

19. P. E. Christensen, H. Schmidt, H. O. Bang, V. Andersen, B. Jordal, O. Jensen, An epidemic of measles in southern Greenland, 1951; measles in virgin soil. III. Measles and tuberculosis. Acta Med Scand 144, 450–454 (1953).

20. P. E. Christensen, H. Schmidt, H. O. Bang, V. Andersen, B. Jordal, O. Jensen, An epidemic of measles in southern Greenland, 1951; measles in virgin soil. II. The epidemic proper. Acta Med Scand 144, 430–449 (1953).

21. S. Y. Chen, S. Anderson, P. K. Kutty, F. Lugo, M. McDonald, P. A. Rota, I. R. Ortega-Sanchez, K. Komatsu, G. L. Armstrong, R. Sunenshine, J. F. Seward, Health Care-Associated Measles Outbreak in the United States After an Importation: Challenges and Economic Impact. Journal of Infectious Diseases 203, 1517–1525 (2011).

22. G. H. Dayan, I. R. Ortega-Sanchez, C. W. LeBaron, M. P. Quinlisk, I. M. R. Team, The cost of containing one case of measles: The economic impact on the public health infrastructure - Iowa, 2004. Pediatrics 116, E1–E4 (2005).

23. T. Pager, Measles Outbreak: Yeshiva’s Preschool Program Is Closed by New York City Health Officials. (2019). 4/15/2019. https://www.nytimes.com/2019/04/15/nyregion/measles-nyc-yeshiva-closing.html.

24. A. Sanders, NYC health officials close two more Williamsburg yeshivas for failure to show immunization records amid measles outbreak. (2019). 6/13/2019. https://www.nydailynews.com/news/politics/ny-city-closes-williamsburg-brooklyn-yeshivas-measles-outbreak-20190613-jogozcbe65ejtalweq255lwkka-story.html.

25. R. M. Anderson, R. M. May, Infectious diseases of humans: dynamics and control. (Oxford University Press, Oxford, 1991).

26. M. J. Keeling, B. T. Grenfell, Disease extinction and community size: Modeling the persistence of measles. Science 275, 65–67 (1997).

27. P. Beck, S. M. Cohen, J. B. Ukeles, R. Miller, Jewish Community Study of New York: 2011, Geographic Profile. revised ed. New York City: UJA-Federation of New York, (2013).

28. W. Yang, J. Li, J. Shaman, Characteristics of measles epidemics in China (1951-2004) and implications for elimination: A case study of three key locations. Plos Comput Biol 15, e1006806 (2019).

29. B. F. Finkenstädt, B. T. Grenfell, Time series modelling of childhood diseases: a dynamical systems approach. Journal of the Royal Statistical Society: Series C (Applied Statistics) 49, 187–205 (2000).

30. W.-m. Liu, H. W. Hethcote, S. A. Levin, Dynamical behavior of epidemiological models with nonlinear incidence rates. Journal of mathematical biology 25, 359–380 (1987).

31. M. J. Keeling, P. Rohani, in Modeling Infectious Diseases in Humans and Animals. (Princeton University Press, 2008), chap. 5, pp. 155.

32. New York City Department of Health and Mental Hygiene, Health Department Reports Eleven New Cases of Measles in Brooklyn’s Orthodox Jewish Community, Urges On Time Vaccination for All Children, Especially Before Traveling to Israel and Other countries Experiencing Measles Outbreaks. (2018). https://www1.nyc.gov/site/doh/about/press/pr2018/pr091-18.page.

33. New York City Department of Health and Mental Hygiene, Measles Outbreak in Orthodox Jewish Community of Brooklyn Continues to Grow — Health Department Urges Parents to Vaccinate Their Children. (2019). 2/28/19. https://www1.nyc.gov/site/doh/about/press/pr2019/measles-outbreak-now-at-121-cases.page.

34. M. S. Arulampalam, S. Maskell, N. Gordon, T. Clapp, A tutorial on particle filters for online nonlinear/non-Gaussian Bayesian tracking. IEEE Trans. Signal Process. 50, 174–188 (2002).

35. M. D. McKay, R. J. Beckman, W. J. Conover, Comparison of three methods for selecting values of input variables in the analysis of output from a computer code. Technometrics 21, 239–245 (1979).

36. W. Yang, A. Karspeck, J. Shaman, Comparison of filtering methods for the modeling and retrospective forecasting of influenza epidemics. PLoS Comput Biol 10, e1003583 (2014).

